# Rehabilitation your way: A randomized trial comparing home-based and self-managed web versus paper programs to improve arm and hand function after stroke

**DOI:** 10.1101/2025.01.14.25320542

**Authors:** Kelly P. Westlake, Sandy McCombe Waller, Lawrence Magdar, Ruth Akinsolotu, Jean Udo, Jane Burridge, Jill Whitall

## Abstract

**Background:** Home-based, self-managed stroke rehabilitation can complement limited healthcare resources. Computer-based programs offer a potential for rehabilitation without the need for complex technology or direct therapist supervision.

**Objective:** To compare effectiveness of a home-based web rehabilitation program (STRONG) versus a paper exercise program (PEP) for stroke-related upper extremity disability.

**Methods:** In this single-blind randomized controlled trial 43 participants with stroke were stratified by motor ability and motivation, then randomly assigned to either STRONG (n=22) or PEP (n=21). Training consisted of a 6-week intervention phase (at least 1 hour daily, 5 days weekly), followed by a 6-week follow-up phase with optional training. Primary outcomes were Fugl-Meyer Upper Extremity Assessment (FMA) and the streamlined timed Wolf Motor Function Test (WMFT-T). Secondary outcomes Motor Activity Log (MAL), WMFT-Function (WMFT-F), Stroke Motivation Scale, and Self-Efficacy Scale.

**Results:** Neither program showed superiority in FMA or WMFT-T. Within-group FMA improvements were similar (Strong: 1.8 points vs PEP: 2.1 points at post-training; 3.3 vs 1.9 at follow-up). However, STRONG participants reported better arm and hand use in daily activities (MAL) at post-training and follow-up. Additionally, STRONG participants showed higher self-efficacy at post-training, though this difference was not maintained at follow-up.

**Conclusion:** Both programs led to modest motor improvements. While STRONG did not outperform PEP in the primary outcomes, participants reported using their affected arm and hand more in daily activities and higher program continuation after the intervention period. Results suggest that home-based, self-managed rehabilitation combining exercises with daily tasks appears beneficial for stroke survivors post-standard rehabilitation.

Clinicaltrials.gov identifier: NCT03484182

## Introduction

Stroke remains a leading cause of acquired disability worldwide.^1^ While stroke incidence has declined in high-income countries,^2^ factors such as improved survival rates and aging populations contribute to increasing numbers of long-term survivors.^3^ Evidence suggests that the amount of therapy is crucial for recovery.^4^ However, healthcare systems struggle to provide sufficient therapy time to optimize recovery outcomes.

While tele-rehabilitation has emerged as one solution, it requires significant costs and effort from both therapists and patients. Self-managed, home-based strategies present a promising approach, facilitating rehabilitation with minimal professional interaction. A Cochrane review of 14 studies on self-managed rehabilitation reported improvements in quality of life and self-efficacy but found no substantial benefits for motor activity,^7^ emphasizing the need for better research into home-based approaches.

We explored “LifeCIT,” a wed-based, individualized, home-based, and self-managed rehabilitation approach, initially trialed in England.^8^ Founded on Self-Determination Theory,^9^ LifeCIT emphasizes motivation and self-efficacy through goal-setting and self-monitoring.^10^ A feasibility trial comparing LifeCIT to usual care demonstrated promise among patients with stroke.^11^ Our program, “STRONG” (STroke Rehabilitation ONline Guide), adapted LifeCIT by incorporating activities for a broader range of stroke impairments, including both bilateral and unilateral training, as well as modifying language and activities for use in the United States.

The purpose of this pragmatic randomized clinical trial was to compare STRONG with a paper-based exercise program (PEP) (also home-based and self-managed) over six-weeks in a randomized controlled trial involving stroke survivors who have completed outpatient rehabilitation. We hypothesized that six weeks of STRONG would lead to clinically meaningful and statistically significant improvements in upper extremity function, both immediately following the intervention (primary endpoint) and six weeks later (secondary endpoint). Additionally, we hypothesized that STRONG would yield statistically significant gains in motivation to train and self-efficacy compared to PEP, both immediately after the intervention and six weeks later.

## Methods

The study adhered to the University Institutional Review Board (IRB), and written informed consent was obtained from all participants. Participants were recruited if they were in the sub-acute (3-6 months) or chronic (> 6 months) phase post-stroke. Initial recruitment was conducted in-person at three local hospitals located in suburban, urban, and rural areas. Recruitment, eligibility assessment, consent, and evaluations shifted online during the COVID-19 pandemic, expanding to include chronic participants nationwide through stroke survivor organizations and online research platforms.

Inclusion criteria were: age ≥30 years; diagnosed with ischemic or hemorrhagic stroke ≥3 months prior to study initiation; discharged from all upper extremity rehabilitation; observable upper extremity impairment; able to move hand forward ≥3 inches from a position of 90°elbow flexion and neutral shoulder; computer/internet access with a video camera for virtual communication and availability of a second person to assist with camera angles during the three assessment sessions (after March 2020); and capable of following instructions provided during the initial screening session. Exclusion criteria included medical, orthopedic, neurological, or cardiovascular conditions that could jeopardize participation; cerebellar stroke; and recent (<3 months) upper extremity botulinum toxin injection.

Sample size calculations were based on the two-arm parallel group pilot RCT (comparing LifeCIT with usual care) (Burridge et al., 2017; Hughes et al. 2017) using the between-group comparison of Wolf Motor Function Test (WMFT) assuming a standard deviation of 0.885 and clinically important effect size of 0.3. Power was set to 80% and the significance level to 5%. These calculations indicated that a total sample size of n= 134 (67 per arm) would be needed.

The randomization schedule, designed by the study statistician (LM), utilized computer-generated pseudo-random numbers with variable block sizes and a 1:1 allocation ratio, stratifying by motor performance and self-efficacy. High motor performance was defined by the ability to independently lift a can with the paretic hand, while high self-efficacy was determined based on an above average score on the Short Self-Efficacy for Exercise Test for Stroke.^12,13^ Participants were randomized after baseline assessment. Research assistants conducting the assessments remained blinded during post-training and follow-up testing.

Both STRONG and PEP groups participated in 6 weeks of training (minimum 1 hour daily, 5 days weekly), with flexible participant-set scheduling options. Participants tracked their training through daily log sheets and received brief (<5mins) weekly check-in phone calls. After post-training evaluation, participants could maintain their respective computer-based or paper-based training activities if they chose to do so for six additional weeks with one mid-period check in phone call.

The STRONG program utilized a web-based computer interface to guide training activities throughout the intervention. Participants first answered questions regarding their abilities on 10 common tasks to determine their functional level. *Level 1* participants used their whole paretic arm as a stabilizer during unilateral tasks (e.g., holding newspaper under arm) or bilateral tasks (e.g., holding paper still while writing with the non-paretic arm). *Level 2* participants could stabilize objects using their paretic hand during bilateral tasks (e.g., holding a jar or toothpaste tube), but could not use it as a manipulator. *Level 3* participants could manipulate objects with their paretic hand for bilateral tasks (e.g., taking a lid from a jar) and unilateral tasks (e.g., lifting and drinking from a can). *Level 4* participants could perform fine-motor activities (e.g., texting and writing) or higher function unilateral tasks (e.g., throwing and catching).

After determining their functional level, STRONG participants chose daily activities from level-appropriate lists. Level 4 participants could select from the Level 3 list or create their own list of meaningful tasks. Activities were categorized into personal care, home chores, or recreational activities and, although the number of activities was self-selected, initial guidance was to select 6 (2 from each category). See Figure 1 for examples of activities for each level in the home chores category. The computer program also included optional training components beyond the 1-hour minimum daily task practice: 5 paretic arm unilateral and bilateral exercises and 5 computer games. The 5 exercises, designed by a physical therapist (SMW), were displayed through images and descriptions and were tailored to each functional level. The number of repetitions for the exercises was self-selected although at least 5 repetitions of each was suggested. The 5 computer games were designed to encourage use of their paretic arm/hand to control the mouse. Again, the time spent on the games was self-selected. Participants were then reminded by the program to log in daily to select their daily practice tasks and again in the evening to report the tasks they had completed throughout the day. The program reassessed functional levels weekly, allowing adjustments to the selection of daily activities, computer-based exercises, and computer games.

**Figure 1.**
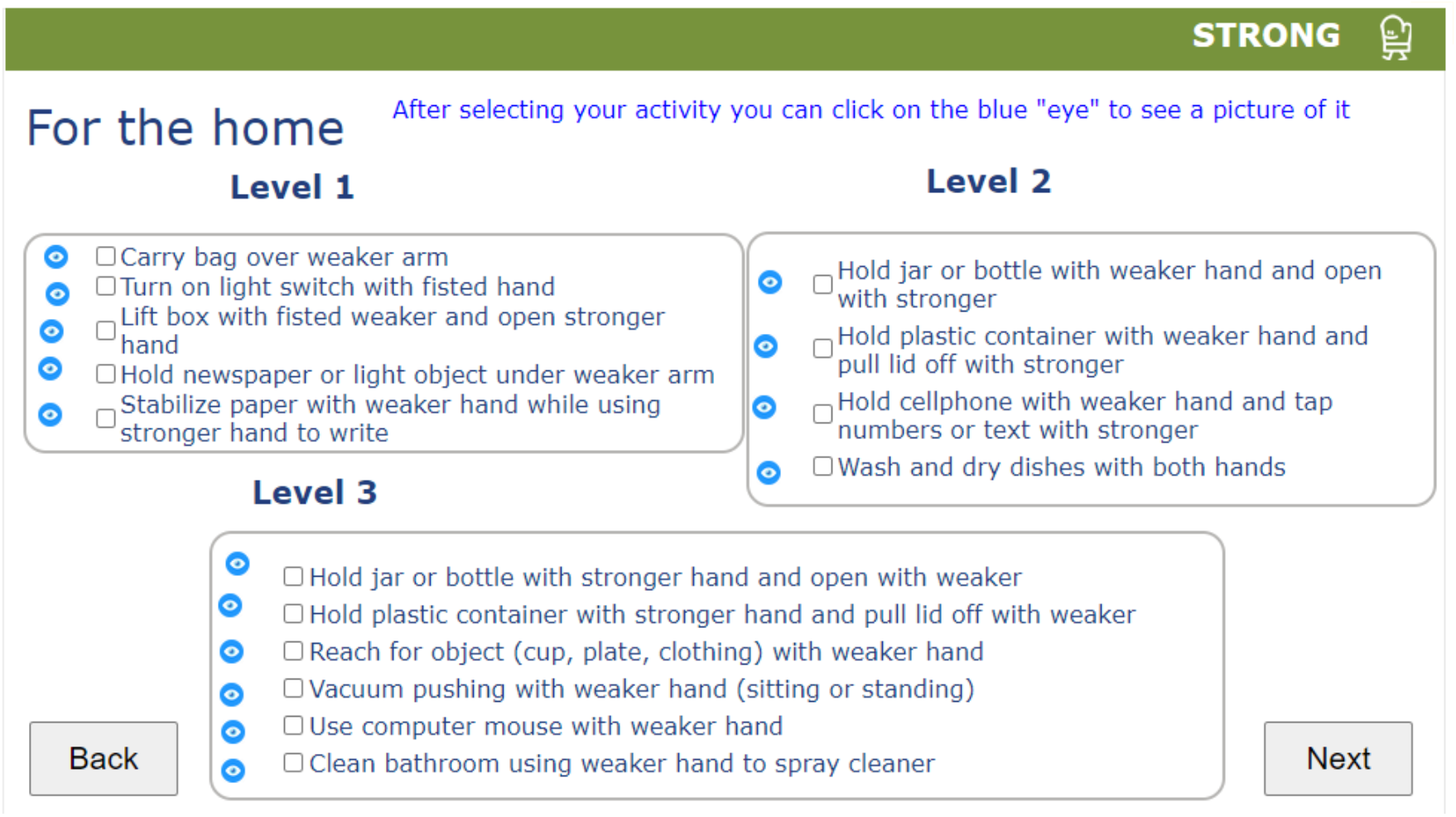
Screenshot of STRONG computer interface for selecting daily practice activities at three levels of motor function ability. Example shown is for home chores. Similar boxes were displayed for personal care and recreational activities. Each day, participants checked the activities they were committed to practicing for a minimum of 1 hour that day within their pre-determined functional level (based on a weekly computer program assessment). Following activity selection for personal care, home chores, and recreational activities, participants were able to optionally complete exercises and video games to promote paretic arm and hand use. At the end of each day, participants logged back in and were prompted to report on the activities and time practiced that day.

PEP featured printed handouts with images and descriptions of the same 5 exercises per functional level that STRONG offered as optional computer-based activities. Participants started with Level 1 with progression guidance based on baseline performance. Instructions regarding dosage was to increase the number of repetitions, speed, or the distance of each exercise if they felt able to do so. As an incentive, PEP participants were offered a crossover opportunity to STRONG following the 6-week follow-up assessment.

Evaluations were conducted at baseline, post-training, and 6-week follow-up. Performance-based assessments were video recorded by blinded research assistants, and a separate blinded assessor (SMW) reviewed and scored the recordings later. SMW also trained the research assistants to conduct these assessments, while another team member (JW) trained them on questionnaire administration. Prior to Mar 2020 (pre-COVID), participants were assessed in person (n=2). After Mar 2020, remaining participants were evaluated virtually. For virtual assessments, participants were asked to gather specific household items from a list (with sizes where applicable) and asked to store these items in a box and use the same ones for each testing session. Additionally, a template for the Wolf Motor Function Test was sent to each participant.

Primary outcomes included the Fugl-Meyer Upper Extremity Assessment (FMA)^14^ and the Six-Item Wolf Motor Function Test-Time (WMFT-T).^15^ Due to virtual data collection constraints, FMA reflex data (three items totaling six points) were not collected. Prior research has demonstrated that excluding reflex data does not compromise the construct validity of the test,^16,17^ resulting in a reduced scoring range of 0 to 60. For the WMFT-T, two tasks were adapted for virtual testing: elbow extension was performed without weight, and turning three cards replaced the key-turning task, as it also involves forearm supination and is validated in the 15-item test ^18^. To test the reliability of assessing the filmed primary testing outcomes, we asked our blinded tester to re-assess 10 completed testing sessions across a variety of participant motor ability levels. For the FMA, the individual item rating agreement was 98.7%. Only two sets of the ten sessions varied by 1 point on the total FM scoring. For the WMFT-timing the average absolute error between measured repeated timing was 0.129 sec or 129 milliseconds with a slight tendency for the repeat testing to be faster than the original.

Secondary Motor Outcomes included the Six-Item Wolf Motor Function Test-Function (WMFT-F) and the Motor Activity Log (MAL). WMFT-F evaluates the quality of movement in the same tasks used for the WMFT-T. The MAL is a patient-reported measure of upper limb activity, divided into two scales: “amount of use” (AU) and “quality of movement” (QM).^19,20^

Secondary Behavioral Outcomes included the Stroke Self-Efficacy Questionnaire^21^ and the Stroke Rehabilitation Motivation Scale.^22^

For the statistical analysis, outcome distributions were first examined at each time point using graphical methods to gain insights into their location, spread, and skewness. Participant characteristics were compared across intervention groups for all randomized individuals. For each outcome variable, we estimated the mean outcome at each time point for each group using a mixed effects model with a random effect for subject. To evaluate the effect of each intervention over time and the differences in these effects, we calculated p-values and confidence intervals for selected contrasts. Our primary analyses followed the intention-to-treat principle, while subsequent analyses considered compliance. We report the differences along with their corresponding 95% confidence intervals (CIs) and effect sizes.

## Results

Figure 2 depicts the study participant flow. From 190 potential participants who expressed interest in the study, 43 were randomized into the STRONG (n=22) or PEP (n=21) groups. Completion group sizes were 16 and 15 for STRONG and PEP, respectively, and 12 (STRONG) and 10 (PEP) at the 6-week follow-up testing session.

**Figure 2.**
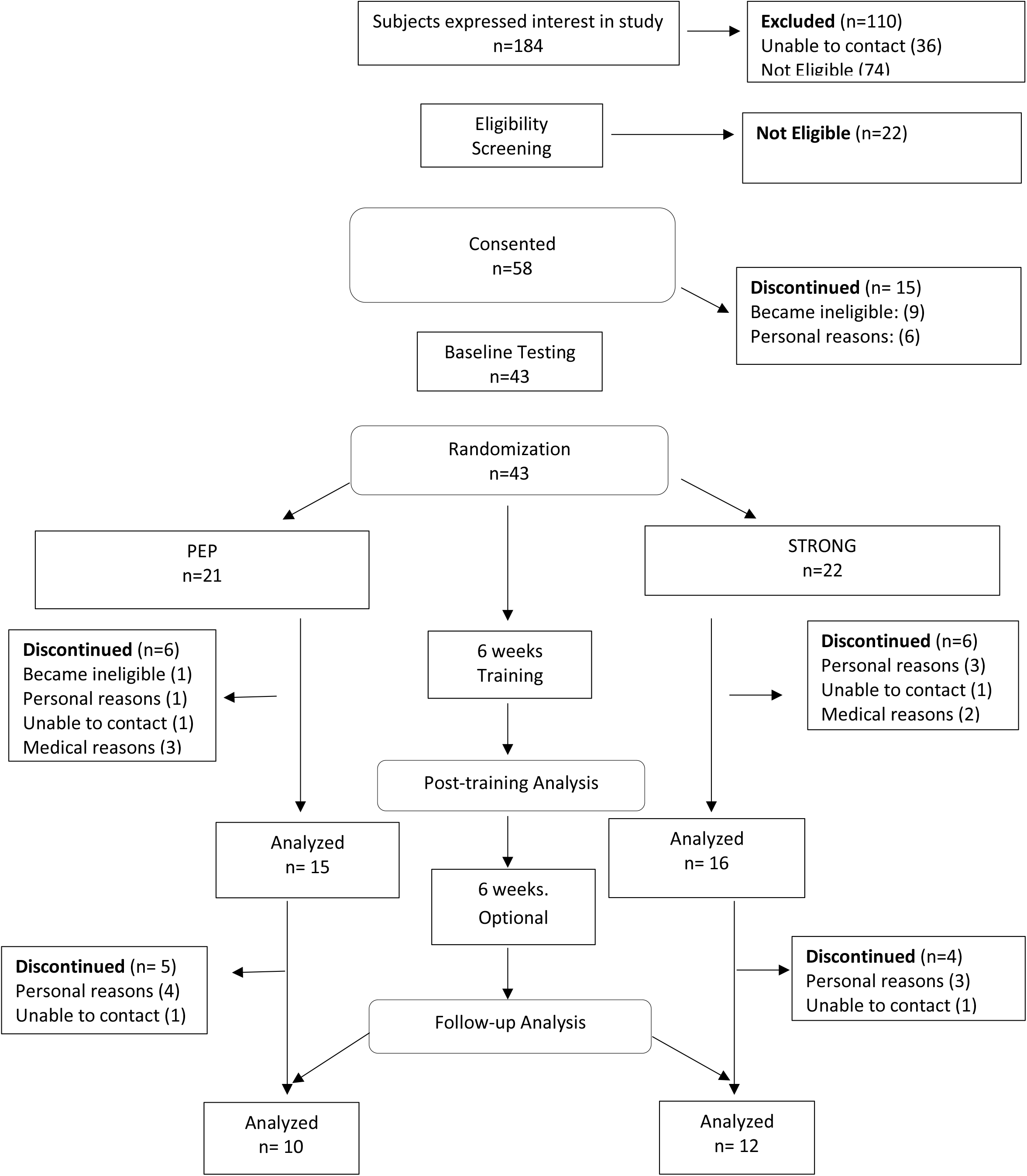
Study participant flow chart

Table 1 presents the demographic baseline characteristics of participants who commenced the study. Randomization procedures revealed no significant differences between the groups regarding age, sex, time since stroke, stroke side or dominance, or baseline functional scores.

**Table 1:** Baseline Characteristics of the Participants.

| Variable | Level | All Patients<br>(n=43) | Strong<br>(n=22) | PEP<br>(n=21) |
| --- | --- | --- | --- | --- |
| Age Group | <40 | 2 ( 5%) | 1 ( 5%) | 1 ( 5%) |
|  | 40-49 | 7 (16%) | 3 (14%) | 4 (19%) |
|  | 50-59 | 16 (37%) | 11 (50%) | 5 (24%) |
|  | 60-69 | 13 (30%) | 5 (23%) | 8 (38%) |
|  | 70+ | 5 (12%) | 2 ( 9%) | 3 (14%) |
| Sex | Female:Male | 22:21 (51%:49%) | 12:10 (55%:45%) | 10:11 (48%:52%) |
| Years Since Stroke | <2 yrs | 10 (23%) | 5 (23%) | 5 (24%) |
|  | 2-4 yrs | 11 (26%) | 7 (32%) | 4 (19%) |
|  | 4-6 yrs | 4 (9%) | 3 (14%) | 1 ( 5%) |
|  | 6+yrs | 9 (21%) | 3 (14%) | 6 (29%) |
|  | Missing | 9 (21%) | 4 (18%) | 5 (24%) |
| Dominant Side | Left:Right | 6:37 (14%:86%) | 3:19 (14%:86%) | 3:18 (14%:86%) |
| Stroke Dominance<br>Side | Motor Dominant | 12 (28%) | 9 (41%) | 3 (14%) |
|  | Motor Nondominant | 31 (72%) | 13 (59%) | 18 (86%) |
| Paretic Side | Left:Right | 27:16 (63%:37%) | 12:10 (55%:45%) | 15:6 (71%:29%) |
| FuglMeyer Total: Mean (SD) Max = 60 |  | 33.71 (17.67) | 33.29 (17.73) | 34.14 (18.04) |
| Wolf Time (secs): Mean (SD) Max = < 1sec |  | 38.97 (41.95) | 42.38 (41.20) | 35.56 (43.43) |
| Wolf Functional: Mean (SD) Max =5 |  | 2.90 ( 1.41) | 2.82 ( 1.40) | 2.98 ( 1.45) |
| MAL Amount of Time: Mean (SD) Max = 5 |  | 1.22 ( 0.97) | 1.05 ( 1.05) | 1.39 ( 0.87) |
| MAL Quality: Mean (SD) Max = 5 |  | 1.38 ( 1.05) | 1.13 ( 1.11) | 1.64 ( 0.95) |
| Self-efficacy: Mean (SD) Max = 141 |  | 96.67 (22.12) | 97.57 (22.50) | 95.76 (22.26) |
| Motivation: Mean (SD) Max = 140 |  | 113.3 ( 8.85) | 115.7 ( 8.16) | 111.0 ( 9.09) |

HEP = home exercise program; SD = standard deviation; MAL = Movement Activity Log; All measured items are scored from low to high with high being best except Wolf Time where a low score is a better performance and 120 seconds is the worst possible score.

Results for all outcome variables are detailed in Table 2, which includes primary motor outcomes followed by secondary motor and behavioral outcomes.

**Table 2.** Model-based estimates of mean outcomes by group at each time point based on a mixed effects longitudinal regression model.

| Outcome | Group | Baseline<br>Mean<br>(SE) | Post-training<br>Mean (SE) | Mean<br>Change from<br>baseline<br>(95% CI) | P-value<br>within-group | Mean<br>Difference<br>between<br>groups in<br>change<br>(95% CI) | P-value<br>between<br>groups | Follow-up<br>Mean<br>(SE) | Change<br>from<br>baseline | P-value<br>within-group | Difference<br>(95% CI) | P-value<br>between<br>groups |
| --- | --- | --- | --- | --- | --- | --- | --- | --- | --- | --- | --- | --- |
| FMA | STRONG | 33.7 (2.7) | 35.5 (2.8) | 1.8 (0.3, 3.4) | 0.023* | -0.3<br>(-2.6, 1.9) | 0.76 | 37.0 (2.9) | 3.3 (1.4, 5.1) | <0.001* | 1.3<br>(-1.3, 4.0) | 0.31 |
|  | PEP |  | 35.9 (2.8) | 2.1 (0.5, 3.8) | 0.014* |  |  | 35.6 (2.9) | 1.9 (0.1, 3.8) | 0.044* |  |  |
| WMFT-T | STRONG | 39.0 (6.6) | 37.6 (7.0) | -1.3 (6.7,4.0) | 0.62 | -1.4<br>(-9.3, 6.6) | 0.73 | 33.9 (7.2) | -5.1<br>(-11.6,1.4) | 0.12 | -7.3<br>(-16.4, 1.9) | 0.12 |
|  | PEP |  | 39.0 (7.1) | 0.0 (-5.9,5.9) | 0.99 |  |  | 41.1 (7.2) | 2.1 (-4.4,8.7) | 0.51 |  |  |
| WMFT- F | STRONG | 2.9 (0.2) | 3.1 (0.2) | 0.2 (0.0, 0.4) | 0.017* | 0.1<br>(-0.1, 0.4) | 0.32 | 3.0 (0.2) | 0.1 (-0.1,0.3) | 0.28 | -0.1<br>(-0.4, 0.2) | 0.43 |
|  | PEP |  | 3.0 (0.2) | 0.1 (-0.1,0.3) | 0.38 |  |  | 3.1 (0.2) | 0.2 (0.0, 0.4) | 0.034* |  |  |
| MAL<br>Amount | STRONG | 1.2 (0.2) | 2.3 (0.2) | 1.1 (0.7, 1.5) | <0.001* | 0.6<br>(0.1, 1.1) | 0.013* | 2.3 (0.2) | 1.1 (0.7, 1.5) | <0.001* | 0.7<br>(0.1, 1.2) | 0.024* |
|  | PEP |  | 1.7 (0.2) | 0.5 (0.1, 0.8) | 0.015* |  |  | 1.7 (0.2) | 0.4 (0.0, 0.9) | 0.045* |  |  |
| MAL<br>Quality | STRONG | 1.4 (0.2) | 2.4 (0.2) | 1.0 (0.6,1.4) | <0.001* | 0.7<br>(0.2, 1.2) | 0.011* | 2.4 (0.2) | 1.0 (0.6, 1.4) | <0.001* | 0.7<br>(0.1, 1.2) | 0.023* |
|  | PEP |  | 1.7 (0.2) | 0.3 (-0.1,0.7) | 0.094 |  |  | 1.7 (0.2) | 0.3 (-0.7,0.7) | 0.15 |  |  |
| Self-Efficacy | STRONG | 96.7 (3.4) | 102.9 (4.1) | 6.2(0.2,12.2) | 0.043* | 7.7<br>(-0.8,16.2) | 0.075 | 101.2(4.3) | 4.5(2.2,11.3) | 0.18 | 6.4<br>(-3.4, 16.3) | 0.19 |
|  | PEP |  | 95.2 (4.1) | -1.5(-7.7,4.7) | 0.64 |  |  | 94.8 (4.6) | -1.9 (-9.2,5.4) | 0.60 |  |  |
| Motiv | STRONG | 113.3<br>(1.4) | 109.1 (2.0) | -4.2(-7.9,-.5) | 0.027* | 0.3<br>(-4.8, 5.4) | 0.90 | 111.4(2.2) | -1.9 (-6.1,2.3) | 0.37 | 2.5<br>(-3.5, 8.4) | 0.41 |
|  | PEP |  | 108.8 (2.0) | -4.5(-8.4,-.7) | 0.021* |  |  | 109.0(2.3) | -4.4 (-8.9, 0.1) | 0.057 |  |  |

\*p<0.05

### Primary Outcomes

At post-training, both intervention groups demonstrated improvements in the primary motor outcome of FMA scores, with no significant between-group differences. These improvements persisted at the 6-week follow-up assessment. Neither group showed significant WMFT-T improvements, although STRONG showed a non-significant trend towards improvements (39.0s to 33.9s) compared to a slightly increased time in the PEP group (39.0s to 41.1s).

### Secondary Motor Outcomes

At post-training, the STRONG group demonstrated significantly improved WMFT-F, while PEP showed no improvement and there was no between-group difference. Improvements in the STRONG group were not maintained at the 6-week follow-up assessment. For the self-reported measures, the STRONG group demonstrated significantly greater improvements in daily arm use (MAL-A) and movement quality (MAL-Q) compared to PEP, with these group differences maintained at follow-up.

### Secondary Behavioral Outcomes

At post-training, only the STRONG group demonstrated a significant within-group improvement in Self-Efficacy, although this improvement was not maintained at the 6-week follow-up. Regarding the Motivation Scale, both groups experienced a decline in motivation at the post-training assessment, recovering near baseline at follow-up.

Adherence to the goal of 30 hours of training over 6 weeks was generally good among participants who reported their training times. On average, STRONG participants (n=9) logged 33.4 hours (SD 14.8; range 17-58.3), versus PEP participants (n=7) 26.4 hours (SD 7.8; range 14.8-37). Adherence varied, with only one PEP participant exceeding the target compared to three from STRONG. Two caveats must be noted: first, daily logs were introduced for participants starting with participant #14, resulting in five prior participants (3 PEP and 2 STRONG) not having logs to complete. Second, five participants from each group did not return their record sheets despite reminders, leading to an overall log return rate of 62%.

Willingness to continue training during the follow up period was notably higher among STRONG participants, with five out of seven completers continuing training compared to none from PEP. Additionally, interest in the STRONG program was evident, as five out of ten PEP participants who completed the study expressed a desire to cross over to STRONG, while only one of twelve STRONG participants chose to transition to PEP.

There were no serious adverse events reported among participants in either the STRONG or PEP group.

## Discussion

In this randomized clinical trial, we compared two home-based, self-managed upper extremity rehabilitation programs (STRONG and PEP). Assessments were conducted immediately post training (6-weeks) and at a follow-up, six weeks post-training completion. Participants were allowed to continue with their intervention after the 6-week assessment. This pragmatic clinically meaningful option generated useful data on acceptance of each intervention. Our primary hypothesis was that the web-based activity/exercise program (STRONG) would outperform the paper-based exercise program (PEP) at the primary endpoint after 6 weeks of training. This hypothesis of STRONG superiority was not supported for the primary outcomes, but both interventions showed sustained within-group FMA improvements. In terms of secondary outcomes, STRONG demonstrated several advantages, with participants reporting greater daily engagement of the paretic arm and improved movement quality compared to the PEP group. Additionally, only STRONG participants exhibited improvements in self-efficacy following training.

The findings for FMA align with previous studies showing comparable FMA improvements in both experimental and control groups with equivalent training doses, as reported in a comprehensive systematic review with similar results in 69% of the analyzed studies.^23^ Importantly, our results suggest that modest improvements in FMA can be achieved even with a home-based, self-managed intervention consisting solely of written exercises.

The absence of significant changes in the WMFT-T was unexpected, given its prior sensitivity to change resulting from lab-based upper extremity rehabilitation.^26–29^ A contributing factor to the overall lack of improvement in WMFT-T may be that neither the STRONG nor PEP groups were specifically instructed to prioritize speed during their training. Our focus was on ensuring participants engaged in the activities for a set duration. Although we did not find statistical differences between groups in this outcome, the confidence interval for the sustained reduction in WMFT-T suggested the potential for a greater reduction of up to 16.4 seconds in the STRONG group.

Encouragingly, STRONG is superior to PEP based on secondary motor and behavioral outcomes. STRONG participants reported greater engagement in activities of daily living and improved movement quality compared to PEP participants, with these gains sustained after training became optional. Notably, improvements in the STRONG group reached the MCID for the MAL in acute/subacute populations.^30^ STRONG’s effectiveness likely stemmed from its emphasis on meaningful activities, built-in weekly self-assessments, and quantitative feedback system. The program’s focus on daily living tasks over generic exercises enhanced MAL scores while promoting participant autonomy and engagement. Moreover, only after STRONG training did we observe enhancements in the functional scores of the WMFT-F. While these improvements were not maintained at the six-week follow-up, they suggest that STRONG may be more effective in inducing changes in motor function compared to PEP.

The observation that PEP participants showed within-group sustained improvement in the quantity and a temporary enhancement in the quality of their activities of daily living, relative to baseline measurements, is noteworthy, given that their training was not task specific. This outcome suggests a successful transfer of functional skills to tasks beyond those practiced. It is possible that the exercises selected for both groups facilitated impairment-based improvements, as evidenced by the FMA results, which were generalizable to tasks of daily living. Given that the STRONG protocol allows for the practice of exercises alongside specific tasks, we tentatively propose that this combined approach, with a greater emphasis on task practice, may be more beneficial than focusing solely on either tasks or exercises.

Overall, our participants demonstrated high motivation at baseline, likely reflecting their commitment to a 12-week study without compensation, which necessitated considerable self-discipline to manage their interventions. However, both groups exhibited a decline in motivation scores over the training period. This decline may suggest that the training experience had a demotivating effect, although it does not account for the simultaneous increase in self-efficacy observed in the STRONG group. It is more plausible that the motivation questionnaire captured a distinct motivational construct to that which contributed to adherence to the training regimen for both groups. Additionally, the design of the questionnaire for the sub-acute population may have influenced these results.

The study design possessed several strengths, including a pragmatic approach that allowed participants to choose their practice times and intensity beyond the suggested minimum. Additionally, the provision of an alternative intervention rather than a standard care control and the inclusion of a follow-up assessment enhanced the study’s robustness. Both groups received equal amounts of information and interaction from the research team, thereby increasing the internal validity of the comparisons.

Key limitations include reduced sample size and heterogeneous population mixing. We were unable to recruit the intended sample size. Nevertheless, the study maintained sufficient statistical power to detect between-group differences in a secondary motor task (MAL) and one of the behavioral measures. A related design limitation was the failure to restrict participants to those recently discharged from outpatient rehabilitation (the sub-acute population), which was the original aim. A more homogeneous sub-acute cohort would have minimized variability due to motivation and proximity to the stroke, thereby enhancing internal validity.

The promising results from both, particularly STRONG, home-based programs suggest effectiveness for post-stroke upper extremity rehabilitation. Both our interventions performed favorably. These findings support computer-based programs for improving home-based practice and function, and next iterations of STRONG will require technological enhancements to improve compliance assessment. Future steps include developing a comprehensive Level 3 randomized controlled trial, incorporating additional methods to enhance patient engagement.

## Conclusions

Both STRONG and PEP modestly improved motor function, with STRONG showing sustained clinically important advantages in self-reported activity amount and quality. STRONG uniquely enhanced self-efficacy and demonstrated superior participant retention of ongoing use of the STRONG program even when exercising became optional. These findings support home-based, self-managed rehabilitation potential, particularly incorporating web-based interfaces, participant choice, and combined daily task activities and video game-based exercises.

## Data Availability

All data produced in the present study are available upon reasonable request to the authors

## Acknowledgements and Funding

The study was supported by NIDILRR Field Initiated Grant 90IFRE0011.

The authors thank all of the participants and their carer in an assistive role and Torran Claiborne, Ashley Graham, Nesreen Alissa, Sebastian Pollett and Sara Demain who contributed to the research. We also thank Kitty Poole, Phil Worthington and Kenneth Wittington who were our stroke advisory consultants throughout the grant.

